# Death Incidents Following Transcatheter Edge-to-Edge Repair (TEER) With the MitraClip Device: Analysis of 10-years Post-marketing Death Reports from the Manufacturer and User Facility Device Experience (MAUDE) Registry

**DOI:** 10.1101/2024.01.14.24301281

**Authors:** Xing-he Sun, Yi-fei Zhao, Yan Li

## Abstract

**Background:** The MitraClip device (Abbott), an FDA-approved transcatheter apparatus, is employed for the management of mitral regurgitation (MR) among patients deemed at high risk for surgical interventions. We analyzed reports in the FDA MAUDE database to evaluate the safety trend in fatal complications associated with MitraClip implantation in the last decade.

**Methods:** We searched the publicly accessible Manufacturer and User Facility Device Experience (MAUDE) database for reports of deaths and injuries associated with MitraClip implantation from October 2013 to September 2023. Duplicate reports were excluded. The Cochran-Armitage test was used to analyze the trend in proportion of fatal events.

**Results:** In the 10 year period after FDA approval, a total of 927 death reports and 9211 injury reports were recorded by the MAUDE database. 592 death reports were included for analysis after excluding duplicates and reports from other sources. The most frequently reported complications were MR (26.69%), tissue damage (24.16%) and hypotension (22.13% ). The most common device-related problems were incomplete coaptation (14.70%), difficult to remove (6.42%), and failure to adhere or bond/positioning failure(4.90%). Most deaths (76.94%) occurred within one year of implantation. The proportion of reported fatal events showed a significant declining trend from 15.9% in 2014-2015 to 3.5% in 2020-2021 (p<0.0001).

**Conclusions:** Analysis of the MAUDE database suggests a decline in reported fatal events associated with MitraClip implantation over time, which may reflect increasing experience with the device. However, passive reporting has limitations including underreporting and incomplete data. Further studies are needed to better characterize the safety profile and complication rates of MitraClip implantation in real-world practice.

## Introduction

Mitral regurgitation (MR) is a pervasive valvular heart condition globally^[1]^, affecting 2-3% of the population^[2]^. In patients aged 50 years or above, the annual mortality rates for moderate primary regurgitation with solely medical treatment reach approximately 3%; for severe cases, it is around 6% ^[3]^. Standard treatment for symptomatic severe MR typically involves surgical repair or replacement, which can potentially enhance patient outcomes. ^[4-6]^. Intervention is suggested in patients who have symptoms indicating severe MR, as well as asymptomatic patients with severe MR and left ventricle systolic dysfunction (class 1B)^[7]^. However, patients with high surgical risk or who cannot undergo surgical treatment have limited therapeutic options. To address this untreated population, the transcatheter edge-to-edge repair (TEER) procedure using the MitraClip® device (Abbott, Menlo Park, CA, USA) was introduced in 2003 and demonstrated effectiveness in reducing mortality^[8-10]^. In the year 2013, the FDA authorization for MitraClip was granted with the purpose of mitigating mitral regurgitation (MR) in patients presenting significant symptomatic MR alongside heart failure symptoms arising from abnormalities in the mitral valve (often referred to as primary or degenerative MR). This intervention was specifically indicated for situations wherein the potential hazards associated with surgery were deemed excessive. In 2019, a novel indication was granted approval, broadening the scope to include the treatment of individuals exhibiting heart failure symptoms as a result of left heart enlargement and diminished function (referred to as secondary or functional MR), alongside moderate-to-severe or severe MR, despite receiving optimal medical therapy, and with structurally normal mitral valves. Nowadays, guidelines recommend using TEER in severely symptomatic patients (NYHA class III or IV) with primary severe MR and a surgical risk that is either high or prohibitive, granted that the mitral valve anatomy permits favorable repair outcomes and the patient has a projected life expectancy of at least 1 year (class 2A)^[7]^. MitraClip presently stands as the first and most widely used percutaneous repair device approved in the United States to treat MR. Ever since its initial use in 2003, MitraClip has been subject to ongoing enhancements aimed at improving device management and reducing the occurrence of complications. The latest iteration of MitraClip, known as “Generation 4,” was introduced in 2019 and boasts a range of improvements. With four unique sizes and an expanded grasping area that is 50% larger, Generation 4 enables implanters to address complex lesions with increased flexibility while minimizing stress on the leaflets. This newest version also introduces innovative features such as side-specific grasping and real-time monitoring of left atrial pressure. Moreover, the preparation process has been simplified, leading to more precise steering^[11]^.

Important data on safety and death has been published since the United State Food and Drug Administration (FDA) approved MitraClip in 2013. Three reports utilizing data from the Manufacturer and User Facility Device Experience (MAUDE) registry of the FDA have been identified. Among them, Galper B Z et al. employed natural language processing to compare event rates of TAVR (transcatheter aortic valve replacement) and MitraClip in the TVT (Transcatheter Valve Therapy) registry, as well as MAUDE data. The event rates obtained from the MAUDE database exhibited no statistical disparities when compared to those reported in the TVT registry ^[12]^. Mahabir et al. demonstrated that median duration between death and reporting was 26 days (range,23-58 days), with 122 (61%) reported within one month. Device-or procedure-related deaths reported in the first four years after the approval of MitraClip accumulated gradually^[13]^. Furthermore, Sheriff N. Dodoo et al. reported^[14]^ that from January 2014 to December 2020, 3370 patients experienced MitraClip related adverse events, with 211 being fatal. An initial ascending pattern in fatal events was observed during the period from 2014 to 2015, which was subsequently followed by a statistically significant decline from 2015 to 2020 (Cochran-Armitage test P = 0.039). With the widespread use of the device and the accumulation of data, we are now able to analyze the evolving safety conditions of TEER using MitraClip in the first decade following FDA approval.

## Material and methods

The FDA administers the MAUDE, a passive surveillance system that requires manufacturers and dealers of medical devices in the United States to report every device-related serious injury, death, or malfunction. Healthcare professionals and patients can also voluntarily report these events^[15]^. The MAUDE registry data is freely accessible to the general public.

Within the database, the search was conducted applying search filters, specifically the product code “NKM” (FDA assigned code for MitraClip) and the product class “Mitral Valve Repair Devices” ^[14]^to retrieve all reports of MitraClip-associated deaths from 1^st^ Oct. 2013(Date of FDA premarket approval^[13]^) to 30^th^ Sep. 2023. The initial two out of the three authors individually interrogated the MAUDE registry and carefully screened for duplicates in the 1061 reports of death. We also searched reports under the “injury” category.

A total of 27 duplicate entries were identified. 300 reports were from research article and 76 from registry data with no unique data. 57 death cases were reported from literature review or meta-analysis with little information provided. There was also 2 reports from study summary, one from before-premarketing clinical trials and 2 from meeting presentation. 4 reported cases were alive. Consequently, a comprehensive analysis was performed on a pool of 592 mortality records. (Figure 1). Of the 9211 injury reports, we excluded 160 death reports, 133 duplicates, 243 research articles, 244 registry reports and 82 supplemental reports. A total of 8349 injury reports were included for death trend analyses.

**Figure 1.**
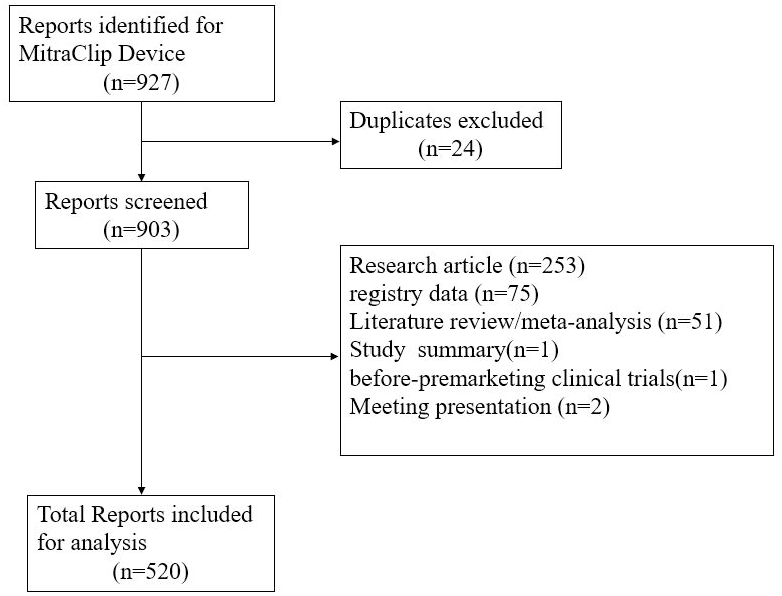
Query of the manufacturer and user facility device experience(MAUDE) database for death reports pertaining to MitraClip implant. Duplicate reports and reports containing inadequate data were excluded from the analysis.

Each narrative death report describes time of report date, device problem, associated complications and comorbidity. Some reports revealed information about time from implantation to death. In order to guarantee the precision of the outcomes, two distinct reviewers independently examined and analyzed all data. Databases that depend on spontaneous reporting, nonetheless, encounter noteworthy restrictions, for instance, the potential for reduced complications and incomplete data. Additionally, the MAUDE database fails to involve the overall count of MitraClip implants conducted. This spontaneous reporting system does not provide mortality rate information. We can only calculate the death rate of reported casualty (death and injury) reports. The primary aim of this study was to observe the trend in the proportion of documented fatal adverse events. Secondary objectives encompassed examining device malfunctions and complications associated with these fatal incidents. To determine a significant trend in the occurrence of fatal adverse events throughout the duration of the study, Cochran-Armitage trend analysis technique was employed. Statistical analysis was conducted using the SAS software (version 9.4, SAS Institute Inc., Cary, NC, USA).

## Results

Among these 592 fatal reports (Figure 2.), 52 types of patients’ problems/complications were identified, potentially leading to death. The 11 most common problems are shown in Figure 3. The top 3 reported complications throughout the study duration were mitral regurgitation (158, 26.69%), tissue damage (143, 24.16%) and hypotension (131,22.13% ).

**Figure 2.**
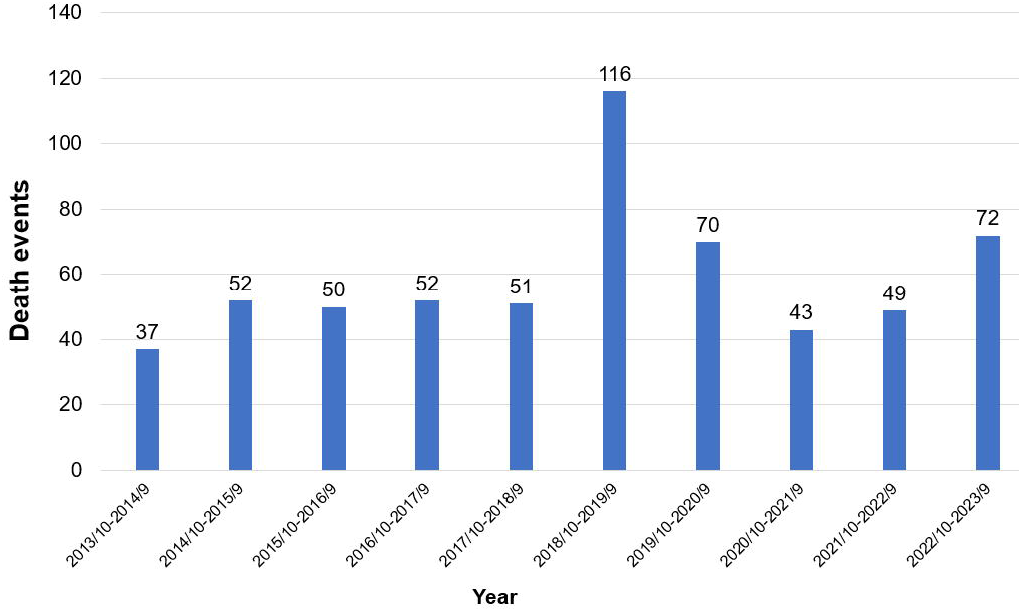
Death events related to MitraClip implant from Oct. 2013 to Sep. 2023. Total count of death events reported each year in 10 years of study duration.

**Figure 3.**
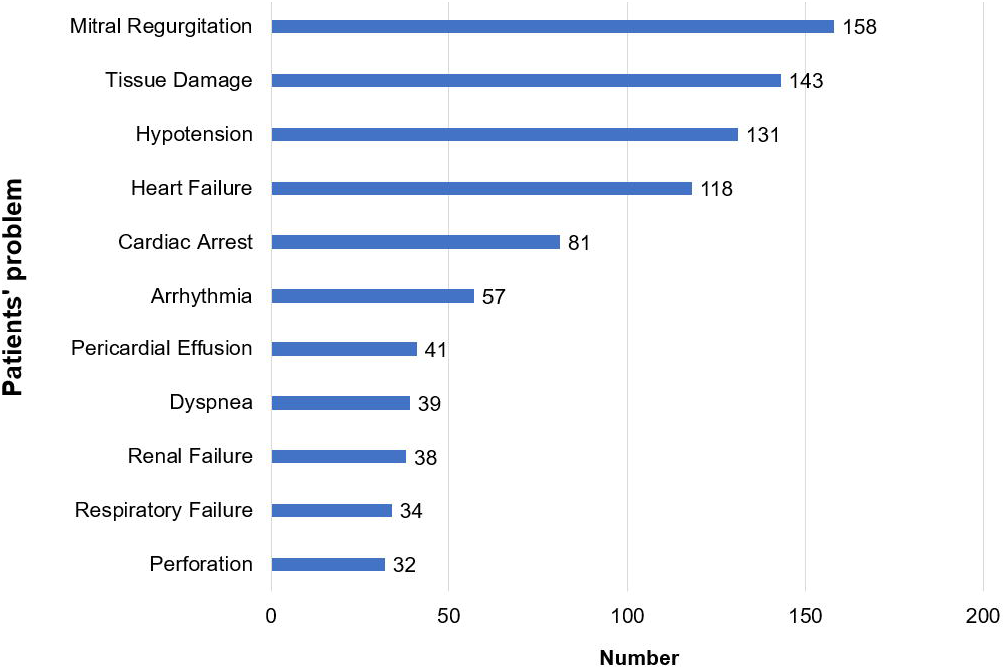
The 11 most common patients’ complications in all the death events associated with MitraClip implant during the study period of tens years(exceeding 30 cases). On the y-axis of the bar chart, various categories of patients’ issues are displayed. The corresponding frequency of occurrences is shown for each category.

Within these 592 fatal reports, we described 31 unique kinds and 394 device problems, considering that certain reports encompassed several device issues. The 11 most common problems exceeding 10 cases are shown in Figure 4. Incomplete coaptation was the most frequently occurring device problem and represented 14.70% of the 592 fatal events (87 events). Difficult to remove (38, 6.42%), failure to adhere or bond and positioning failure(29,4.90%), and improper or incorrect procedure or method(28,4.73%) were all frequently occurring device problems.

**Figure 4.**
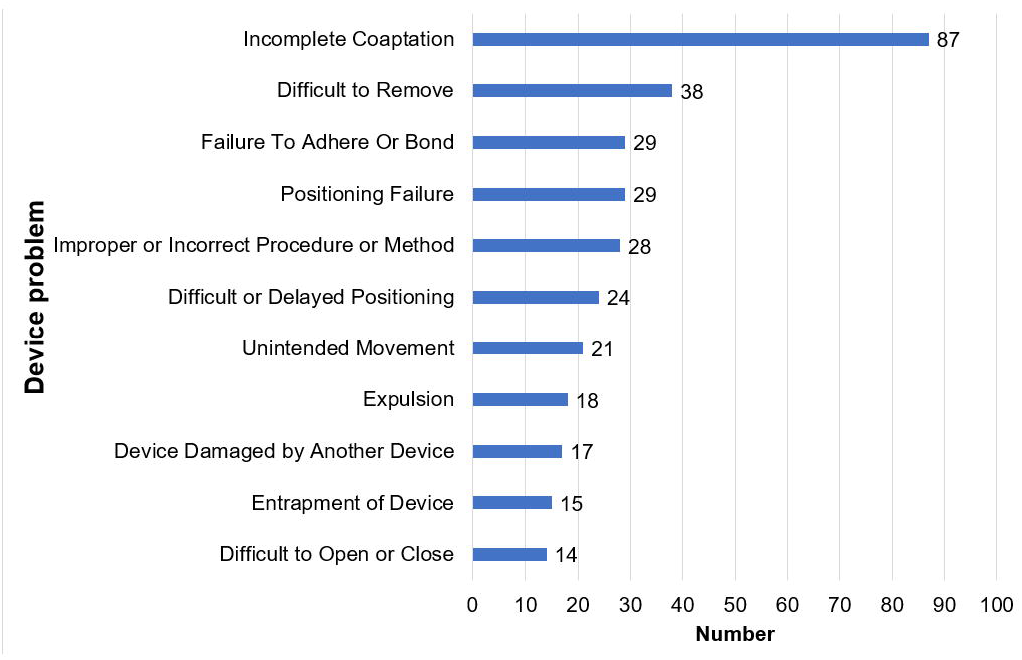
The 11 most common device problems in all the fatal events associated with MitraClip implant during the study period (exceeding 10 cases). On the y-axis of the bar chart, various categories of device problems are displayed. The corresponding frequency of occurrences is shown for each category.

Among the 592 fatal reports, we could speculate on the time from death to the implantation of MitraClip in 490(82.78%) cases. 69(14.08%) of the 490 death reports with enough detail occurred on the same day of implantation. 78 deaths happened in less than one week, 17 in one month, and 377 death reports (76.94%) were described during the first year post MitraClip (Figure 5.). 19(3.21%) of the 592 death reports occurred after the second procedure of implantation. 18 individuals (3.04%) died after mitral valve replacement surgery following unsuccessful implantation of the MitraClip device.

**Figure 5.**
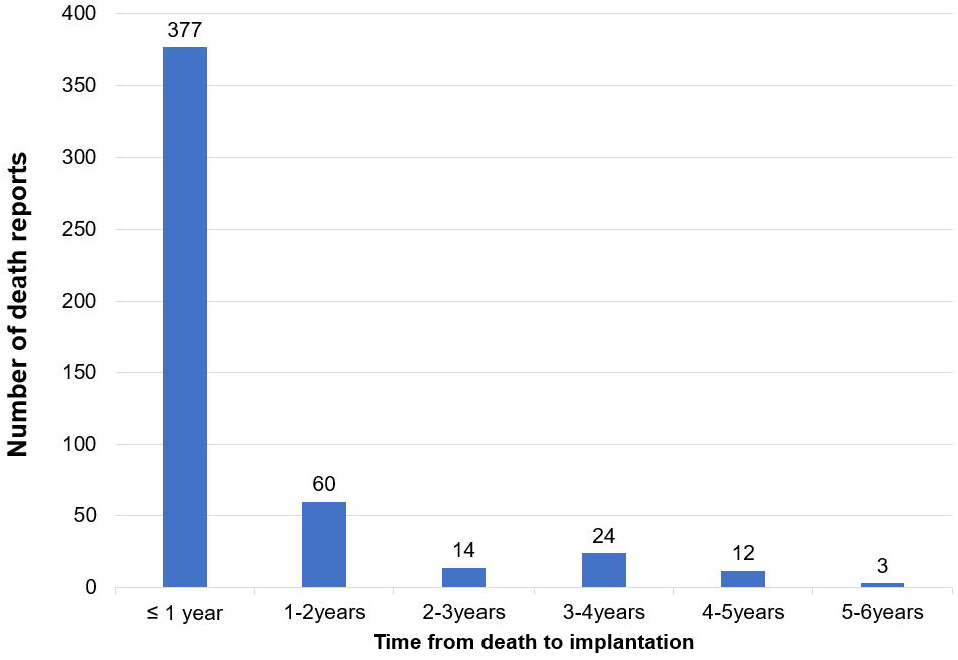
Time from death to implantation of MitraClip of the 490 reports with time detail.

The Cochran-Armitage trend test showed that the proportion of deaths of all casualty events in each year demonstrates a discernible temporal trend that holds statistical significance(*P*<0.0001).

In light of specific numerical values, the mortality composition ratio shows a declining pattern from a peak of 15.9% (n = 52 deaths) in October 2014-September 2015 to a trough of 3.50% (n = 49 deaths) in October 2020-September 2021,with a slightly increase in October 2018-September 2019 (Fig. 6). Casualty events consisted of death reports and device-associated injury reports.

**Figure 6.**
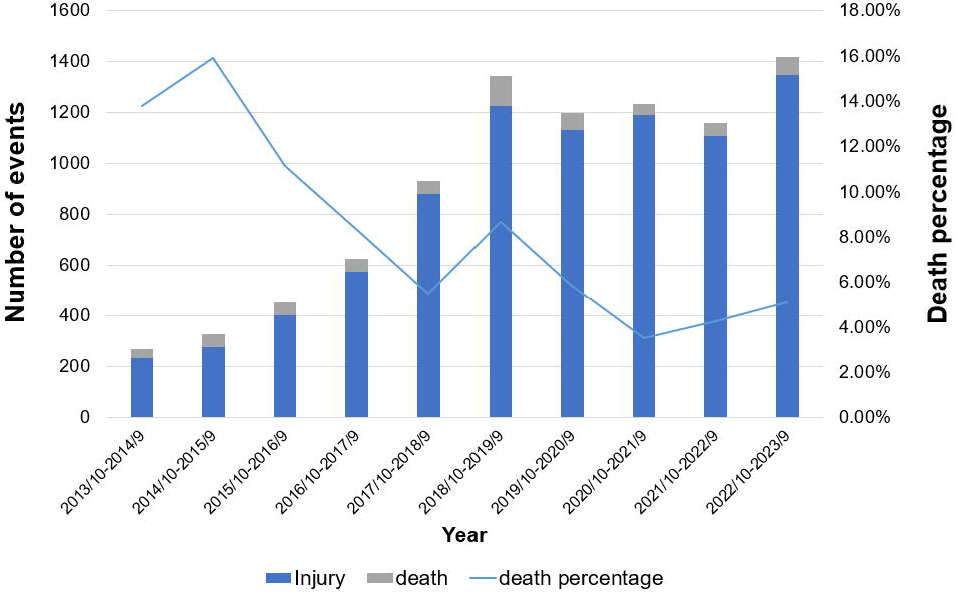
Yearly percentage of death events within all casualty reports in MAUDE database from 2013.10 to 2023.9

## Discussion

This study utilized a nationwide database documenting self-reported adverse events subsequent to the MitraClip procedure. An observable pattern emerged with respect to the reported incidence of mortality. The findings revealed a steady decline in the proportion of reported deaths over all casualty events associated with MitraClip, starting from a peak of 15.9% in 2015 to 3.5% in 2021 over the course of the study duration(*P*<0.0001, employing the Cochran-Armitage test). From 2015 to 2021, there was a reduction of over 80% in the frequency of these reported incidents, indicating a significant decline over the past eight years. The occurrence of fatal events exhibited an initial ascending trajectory between 2014 and 2015, followed by a marked subsequent decline. The observed decline in reported fatalities could potentially indicate an operator and institutional learning curve, possibly attributed to enhanced proficiency in the insertion of these devices, particularly in the later years following their approval, as compared to the initial years.

In a similar vein, the study conducted by Sheriff N. Dodoo et al. ^[14]^ also documented a decline in the occurrence of fatal events between 2015 and 2020 (Cochran-Armitage test *P* = 0.039). The highest proportion of reported death events was observed in 2015, accounting for 44 cases (1.31% of reported adverse events, which encompassed not only fatal events but also device-associated injury reports and device malfunctions). On the contrary, the year 2020 exhibited the lowest percentage of documented fatal events, with a total of 19 cases (constituting merely 0.56% of the reported adverse events). The proportion of death was lower than this study since they used nonfatal events including all injuries and device system malfunction as denominator. In our study, we employed casualty events as the denominator because injury incidents and equipment malfunction incidents may involve a significant degree of overlap and cannot accurately represent the proportion of fatalities in events causing harm to individuals.

Chetaj A. Mahabir’s study^[13]^ involved the analysis of adverse reports obtained from the MAUDE database during the initial four-year period subsequent to FDA approval.21% death happened >1 year post MitraClip, similar with the proportion of 23.06% in this study. The leading three causes of postprocedural mortality were identified as complications necessitating high-risk surgical intervention, detachment or unsuccessful placement of the clip, and harm to the valvular apparatus. However, the top three complications in death reports of this study were incomplete coaptation, difficult to remove and failure to adhere (unsuccessful clip placement).

With the evolution of MitraClip device, adverse complications also changed^[16]^. Among the participants enrolled in the EVEREST II trial, the three most frequently reported non-fatal adverse events within the MitraClip group consisted of blood transfusion, prolonged ventilation exceeding 48 hours, and stroke. The most frequent early phase device complications were single leaflet device attachment and second MitraClip procedure, while mitral valve stenosis was the top late phase device complication^[9]^. In the ACCESS-EU trail^[8]^, renal failure and bleeding were the most frequently occurring adverse events. However, in this study, the top three reasons of death were mitral regurgitation (158, 26.69%), tissue damage (143, 24.16%) and hypotension (131,22.13% ). The presence of the two most significant complications in fatal incidents may potentially indicate underlying engineering deficiencies that can be addressed through subsequent iterations characterized by enhanced precision and refinement.

As for the top device complications, incomplete coaptation might cause mitral regurgitation, the top patient complications of death. It might be caused by improper operation instead of device malfunction. Hence it is imperative for healthcare practitioners to thoroughly review and adhere to the Instructions for Use as well as the recommendations put forth by the device manufacturer. These guidelines encompass crucial information regarding procedural aspects such as implant positioning, locking sequences, clip arm angle establishment, preparation for clip release, and the importance of exercising caution during unlocking of the clip, both during device preparation and throughout the procedure. The collection of voluntary reports concerning adverse events from healthcare providers and device manufacturers plays a pivotal role in the identification and enhanced comprehension of the risks associated with medical devices.

## Limitation

The primary limitations of this study revolve around the inherent characteristics of the data obtained from the MAUDE Registry. Variations in trade product names and company designations can significantly impact the search outcomes. Furthermore, the search process exclusively retrieves records that explicitly include the designated search term(s) provided by the user. Moreover, as the collection of these reports mainly depends on voluntary reports from patients, health care providers and manufacturers, it is likely that certain significant safety events are missing, unreported or reported repeatedly, thereby potentially distorting any findings derived from studies utilizing the MAUDE Registry data. The decline of report over time could be from device not being new with people stop reporting. Additionally, data in the passive surveillance system might be incomplete, in accurate, untimely, or biased. Therefore, it is essential to interpret the results of this study in their appropriate contexts. Moreover, the limitations of this study are further compounded by the absence of comprehensive nationwide records pertaining to the utilization of MitraClip devices. As a result, it becomes impractical to generalize the rates, incidence and prevalence of any given condition solely based on reported events within the MAUDE database. Besides, certain categories of report information, specifically personnel or medical files data such as date of death, date of implantation, and age, are safeguarded under the Freedom of Information Act (FOIA), thus impeding public disclosure. Moreover, the consequence of death is probably the combination of disease itself and the device operation. It is impossible to distinguish from the data. Consequently, confirming the exact cause and timing of a specific event can prove challenging based solely on the information provided within a given report. Lastly, it should be noted that the accuracy of the gathered data remains unverified independently, as the FDA solely collects adverse event reports without authentication.

## Conclusion

This retrospective analysis of the MAUDE Registry has revealed a declined trend in reported fatal events during the study period. It should be acknowledged that due to the limitations inherent in this study and the suboptimal quality of the MAUDE-derived data, definitive attributions regarding this noteworthy finding cannot be drawn. Nevertheless, these results may suggest preliminary evidence of an operator learning curve linked to the use of this particular device. As procedural experience increases among healthcare professionals, there is a likelihood of acquiring valuable expertise in the safe and effective implantation of the MitraClip. Adherence to standardized operating procedures outlined in the manual is also very important in achieving better outcomes. Additionally, ongoing advancements in the engineering of the MitraClip by manufacturers are anticipated to contribute further to its safety profile. However, considering the partial and insufficient quality of the data gathered from the MAUDE Registry, it is imperative to conduct further meticulously designed studies to enhance our comprehension of these documented alterations. This is especially crucial as the clinical effectiveness of the MitraClip device gains more recognition and acceptance.

## Data Availability

All data produced are available online at https://www.accessdata.fda.gov/scripts/cdrh/cfdocs/cfMAUDE/search.CFM

## Notes

The authors report no financial relationships or conflicts of interest regarding the content herein.

### Competing Interest Statement

The authors have declared no competing interest.

### Funding Statement

This study did not receive any funding

### Summary of Updates

Supplemented data from September 2022 to October 2023

## Reference

[1] Nkomo V T, Gardin J M, Skelton T N, et al. Burden of valvular heart diseases: a population-based study[J]. Lancet, 2006,368(9540):1005–1011.

[2] Perlowski A, St G F, Glower D G, et al. Percutanenous therapies for mitral regurgitation[J]. Curr Probl Cardiol, 2012,37(2):42–68.

[3] Enriquez-Sarano M, Akins C W, Vahanian A. Mitral regurgitation[J]. Lancet, 2009,373(9672):1382–1394.

[4] Alegria-Barrero E, Franzen O W. Mitral Regurgitation - A Multidisciplinary Challenge[J]. Eur Cardiol, 2014,9(1):49–53.

[5] Phillips H R, Levine F H, Carter J E, et al. Mitral valve replacement for isolated mitral regurgitation: analysis of clinical course and late postoperative left ventricular ejection fraction[J]. Am J Cardiol, 1981,48(4):647–654.

[6] Ling L H, Enriquez-Sarano M, Seward J B, et al. Clinical outcome of mitral regurgitation due to flail leaflet[J]. N Engl J Med, 1996,335(19):1417–1423.

[7] Otto C M, Nishimura R A, Bonow R O, et al. 2020 ACC/AHA Guideline for the Management of Patients With Valvular Heart Disease: Executive Summary: A Report of the American College of Cardiology/American Heart Association Joint Committee on Clinical Practice Guidelines[J]. Circulation, 2021,143(5):e35–e71.

[8] Maisano F, Franzen O, Baldus S, et al. Percutaneous mitral valve interventions in the real world: early and 1-year results from the ACCESS-EU, a prospective, multicenter, nonrandomized post-approval study of the MitraClip therapy in Europe[J]. J Am Coll Cardiol, 2013,62(12):1052–1061.

[9] Glower D D, Kar S, Trento A, et al. Percutaneous mitral valve repair for mitral regurgitation in high-risk patients: results of the EVEREST II study[J]. J Am Coll Cardiol, 2014,64(2):172–181.

[10] Stone G W, Lindenfeld J, Abraham W T, et al. Transcatheter Mitral-Valve Repair in Patients with Heart Failure[J]. N Engl J Med, 2018,379(24):2307–2318.

[11] Chakravarty T, Makar M, Patel D, et al. Transcatheter Edge-to-Edge Mitral Valve Repair With the MitraClip G4 System[J]. JACC Cardiovasc Interv, 2020,13(20):2402–2414.

[12] Galper B Z, Beery D E, Leighton G, et al. Comparison of adverse event and device problem rates for transcatheter aortic valve replacement and Mitraclip procedures as reported by the Transcatheter Valve Therapy Registry and the Food and Drug Administration postmarket surveillance data[J]. Am Heart J, 2018,198:64–74.

[13] Mahabir C A, DeFilippis E M, Aggarwal S, et al. The First 4 Years of Postmarketing Safety Surveillance Related to the MitraClip Device: A United States Food and Drug Administration MAUDE Experience[J]. J Invasive Cardiol, 2020,32(5):E130–E132.

[14] Dodoo S N, Okoh A K, Oseni A, et al. Adverse Events Following Transcatheter Edge-to-Edge Repair (TEER) Using MitraClip: Lessons Learned From the Manufacturer and User Facility Device Experience (MAUDE) Registry[J]. Cardiovasc Revasc Med, 2022,39:101–105.

[15] MAUDE. Manufacturer and user facility device experience. [EB/OL]. [11 Nov]. https://www.accessdata.fda.gov/scripts/cdrh/cfdocs/cfmaude/search.cfm.

[16] Schnitzler K, Hell M, Geyer M, et al. Complications Following MitraClip Implantation[J]. 2021,23(9):131.

